# Antibody response to symptomatic infection with SARS-CoV-2 Omicron variant viruses, December 2021—June 2022

**DOI:** 10.1101/2023.11.17.23298700

**Authors:** Ryan Sandford, Ruchi Yadav, Emma K. Noble, Kelsey Sumner, Devyani Joshi, Sara Y. Tartof, Karen J. Wernli, Emily T. Martin, Manjusha Gaglani, Richard K. Zimmerman, H. Keipp Talbot, Carlos G. Grijalva, Edward A. Belongia, Christina Carlson, Melissa Coughlin, Brendan Flannery, Brad Pearce, Eric Rogier

**Affiliations:** Centers for Disease Control and Prevention, Atlanta, GA, USA; Oak Ridge Institute for Science and Education, Oak Ridge, TN, USA; Rollins School of Public Health, Atlanta, GA, USA; Kaiser Permanente Southern California, Department of Research & Evaluation; Department of Health Systems Science, Kaiser Permanente Bernard J. Tyson School of Medicine, Pasadena, CA, USA; Kaiser Permanente Washington Health Research Institute, Seattle, WA, USA; University of Michigan School of Public Health, Ann Arbor, MI, USA; Baylor Scott & White Health, Temple, TX, USA; Texas A&M University College of Medicine, Temple, TX, USA; University of Pittsburgh, Pittsburgh, PA, USA; Vanderbilt University Medical Center, Nashville, TN, USA; Marshfield Clinic Research Institute, Marshfield, WI, USA

**Keywords:** COVID-19, SARS-CoV-2 infection, omicron subvariants, immune response

## Abstract

To describe humoral immune responses to symptomatic SARS-CoV-2 infection, we assessed immunoglobulin G binding antibody levels using a commercial multiplex bead assay against SARS-CoV-2 ancestral spike protein receptor binding domain (RBD) and nucleocapsid protein (N). We measured binding antibody units per mL (BAU/mL) during acute illness within 5 days of illness onset and during convalescence in 105 ambulatory patients with laboratory-confirmed SARS-CoV-2 infection with Omicron variant viruses. Comparing acute- to convalescent phase antibody concentrations, geometric mean anti-N antibody concentrations increased 47-fold from 5.5 to 259 BAU/mL. Anti-RBD antibody concentrations increased 2.5-fold from 1258 to 3189 BAU/mL.

## INTRODUCTION

Humoral immune responses to infection with SARS-CoV-2 include production of immunoglobulin G (IgG) antibodies that bind to spike (S) glycoprotein, including the receptor binding domain (RBD) within S, as well as the nucleocapsid (N) protein. Virus neutralization titers and antibodies that bind to RBD and other S protein epitopes have been associated with protection against symptomatic infection with ancestral and pre-Omicron SARS-CoV-2 variants [1-4]. Because antibodies to N protein are not elicited by U.S.-licensed COVID-19 vaccines, the presence of anti-N antibodies can be an indicator of past SARS-CoV-2 infection among vaccinated and unvaccinated individuals [5, 6]. Elevated levels of anti-N antibody may indicate more recent SARS-CoV-2 infection [7]. As new SARS-CoV-2 variants have emerged, serologic assays that quantify IgG antibody binding to spike and nucleocapsid protein antigen have been used to evaluate humoral response to SARS-CoV-2 infection [8]. Post-infection antibodies may reduce risk of re-infection with new SARS-CoV-2 variants, suppress viral replication and reduce COVID-19 disease severity following re-infection [2, 8].

We previously showed that anti-N antibody seropositivity modified COVID-19 mRNA vaccine effectiveness against symptomatic SARS-CoV-2 infection with SARS-CoV-2 Delta and Omicron variants [9]. Among acutely ill patients, higher levels of binding antibodies against ancestral spike protein reduced the odds of testing positive for SARS-CoV-2 Delta and Omicron variants. Here, we assessed humoral immune response to SARS-CoV-2 infection comparing acute- and convalescent-phase IgG antibody levels against N and ancestral spike RBD antigens among case patients infected during Omicron-predominant variant periods from December 2021 through June 2022.

## METHODS

### Study population and sample collection

Between December 2021 and June 2022, respiratory swabs and acute-phase dried blood spots on filter paper were obtained <5 days after symptom onset from ambulatory patients with respiratory illness enrolled in the US Influenza Vaccine Effectiveness Network, as previously described [9, 10]. Patients who tested positive for SARS-CoV-2 by nucleic acid amplification in respiratory specimens were scheduled for convalescent-phase blood sample collection at 21–56 days after enrollment. Data collected from enrolled participants included patient age, date of illness onset, symptoms associated with COVID-like illness, self-reported presence of specified underlying medical conditions, documented COVID-19 vaccination history including dates of COVID-19 vaccination, self-reported laboratory-confirmed COVID-19 <90 or ≥90 days before current illness or electronic medical record of a positive COVID-19 test.

### Serologic assays

Methods for estimating SARS-CoV-2 binding antibody concentration from blood spots have been previously published [11, 12]. Dried blood spot specimens were tested for immunoglobulin G (IgG) antibodies against SARS-CoV-2 recombinant antigens representing ancestral spike protein RBD and nucleocapsid protein using a validated multiplex bead assay (FlexImmArray™ SARS-CoV-2 Human IgG Antibody Test, Tetracore, Rockville, MD) on a Luminex MAGPIX instrument with LX200 flow analyzer (Luminex Corporation, Austin, TX). Eluted specimens were diluted 1:300 and individual specimen median fluorescence intensity (MFI) ratios were calculated compared to antigen-specific human IgG calibrator serum. MFI units were standardized to binding antibody units per mL (BAU/mL) against World Health Organization (WHO) international standards [10, 13].

### Statistical analysis

Antibody concentrations in BAU/mL were log-transformed (log 10) for estimation of geometric mean concentrations (GMC) and 95% confidence intervals (CI) and back-transformed for plots by BAU/mL concentrations. Geometric mean fold rise in bAb concentration was estimated as the geometric mean ratio of bAb concentrations measured at enrollment and follow-up. Fold rise in anti-N and anti-RBD antibody concentrations represented response to acute SARS-CoV-2 infection. Associations between antibody fold rise and patient characteristics, vaccination status, baseline N antibody serostatus, and prior positive COVID-19 test were assessed by t-test or ANOVA. Statistical analysis was performed using R version 4.0.3 (R Foundation for Statistical Computing, Vienna, Austria).

## RESULTS

A total of 105 SARS-CoV-2 positive participants had acute-phase blood specimens collected a median of 2 days (range: 0–5) and convalescent-phase specimens collected a median of 36 days (range: 23–61) after symptom onset. Of 48 SARS-CoV-2 positive specimens with genomic sequencing, 43 (90%) belonged to Omicron BA.1 (n= 5) or BA.2 (n=38) lineages, with five sequences belonging to BA.2.12.1 (n=3), BA.4 (n=1) and BA.5 (n=1) lineages. Among 105 case patients with paired specimens, 36% were aged <40 years, 17% were >65 years, 57% were female and 23% had an underlying health condition (Table 1).

**Table:** Concentrations and fold increase in binding antibodies against SARS-CoV-2 nucleocapsid and ancestral spike receptor binding domain antigens during acute- and convalescent-phase of symptomatic infection during Omicron-predominant variant period.

| | Total<br>N = 105<br>(%) | α-SARS-CoV-2 Nucleocapsid Antibody | | | p-value<sup>1</sup> | α-SARS-CoV-2 Spike Receptor Binding Domain Antibody | | | p-value<sup>1</sup> |
| --- | --- | --- | --- | --- | --- | --- | --- | --- | --- |
|  |  | Acute<br>Geometric Mean Concentration, BAU/mL (95% CI) | Convalescent<br>Geometric Mean Concentration, BAU/mL (95% CI) | Geometric Mean Fold Rise (95% CI) |  | Acute<br>Geometric Mean Concentration, BAU/mL (95% CI) | Convalescent<br>Geometric Mean Concentration, BAU/mL (95% CI) | Geometric Mean Fold Rise (95% CI) |  |
| **Overall** | 105 | 5.5 (4.3, 7.1) | 259.4 (200.6, 335.4) | 47.0 (33.5, 65.9) |  | 1257.8 (923.9, 1712.3) | 3188.5 (2638.7, 3853.0) | 2.5 (1.9, 3.3) |  |
| **Age Group** |  |  |  |  | 0.815 |  |  |  | 0.867 |
| 0–39 years | 38 (36) | 5.0 (3.4, 7.4) | 256.9 (160.2, 412.0) | 51.0 (28.0, 93.0) |  | 1131.0 (618.4, 2068.7) | 2837.9 (2091.0, 3851.5) | 2.5 (1.4, 4.6) |  |
| 40–65 years | 49 (47) | 6.7 (4.3, 10.5) | 279.7 (193.9, 403.3) | 41.8 (24.7, 70.9) |  | 1374.4 (965.9, 1955.6) | 3706.1 (3147.8, 4363.4) | 2.7 (1.9, 3.8) |  |
| > 65 years | 18 (17) | 4.0 (2.9, 5.6) | 215.7 (113.7, 409.3) | 54.1 (27.1, 107.8) |  | 1236.4 (463.3, 3299.5) | 2707.5 (1154.6, 6349.4) | 2.2 (1.3, 3.6) |  |
| **Sex** |  |  |  |  | 0.580 |  |  |  | 0.350 |
| Female | 60 (57) | 5.8 (3.9, 8.5) | 294.8 (224.8, 386.6) | 51.1 (32.8, 79.5) |  | 1080.7 (661.6, 1765.2) | 3058.5 (2304.3, 4059.4) | 2.8 (1.9, 4.1) |  |
| Male | 45 (43) | 5.2 (3.9, 7.0) | 218.7 (134.2, 356.4) | 42.0 (24.4, 72.5) |  | 1539.8 (1124.3, 2108.8) | 3370.6 (2648.0, 4290.3) | 2.2 (1.5, 3.3) |  |
| **Underlying medical condition<sup>2</sup>** |  |  |  |  | 0.914 |  |  |  | 0.770 |
| Yes | 24 (23) | 4.8 (3.0, 7.7) | 222.7 (130.2, 380.8) | 46.6 (24.6, 88.0) |  | 1505.0 (995.7, 2274.6) | 3705.4 (3042.7, 4512.3) | 2.5 (1.6, 3.8) |  |
| No | 76 (72) | 5.6 (4.1, 7.8) | 273.7 (199.5, 375.6) | 48.5 (31.6, 74.4) |  | 1112.0 (742.0, 1666.3) | 2970.4 (2303.4, 3830.4) | 2.7 (1.9, 3.8) |  |
| **COVID-19 vaccination status** |  |  |  |  | 0.525 |  |  |  | <0.001 |
| Unvaccinated | 4 (4) | 9.8 (0.8, 125.0) | 1133.6 (631.7, 2034.2) | 115.5 (6.7, 1998.0) |  | 16.0 (0.2, 1556.6) | 149.0 (2.4, 9353.8) | 9.3 (0.3, 273.1) |  |
| 2 mRNA doses | 14 (13) | 3.9 (1.3, 11.9) | 214.3 (120.5, 380.9) | 55.2 (17.6, 173.2) |  | 414.6 (148.1, 1160.5) | 3268.3 (2293.0, 4658.4) | 7.9 (2.6, 24.3) |  |
| $\geq 3$ mRNA doses | 87 (83) | 5.7 (4.4, 7.3) | 250.0 (186.8, 334.5) | 43.9 (30.4, 63.4) | | 1837.7 (1473.7, 2291.6) | 3656.2 (3190.6, 4189.7) | 2.0 (1.6, 2.5) | |
| **Baseline α-SARS-CoV-2 nucleocapsid antibody serostatus<sup>3</sup>** |  |  |  |  | <0.001 |  |  |  | 0.001 |
| Seropositive | 33 (31) | 24.0 (15.4, 37.4) | 350.5 (198.2, 620.0) | 14.6 (7.0, 30.3) |  | 2065.3 (1211.5, 3520.9) | 2685.7 (1881.4, 3834.1) | 1.3 (0.8, 2.1) |  |
| Seronegative | 72 (69) | 2.8 (2.4, 3.2) | 226.0 (171.8, 297.1) | 80.3 (59.4, 108.5) |  | 1002.0 (689.3, 1456.8) | 3449.4 (2750.9, 4325.3) | 3.4 (2.5, 4.7) |  |
| **Prior COVID-19 positive test** |  |  |  |  | 0.754 |  |  |  | 0.710 |
| Yes | 7 (7) | 33.1 (7.1, 153.8) | 983.4 (545.9, 1771.2) | 29.7 (4.5, 196.5) |  | 1476.3 (326.9, 6665.9) | 2928.8 (1414.9, 6062.7) | 2.0 (0.8, 4.9) |  |
| No | 98 (93) | 4.9 (3.8, 6.2) | 235.8 (181.2, 307.0) | 48.5 (34.3, 68.7) |  | 1243.5 (902.3, 1713.5) | 3207.9 (2628.6, 3914.9) | 2.6 (1.9, 3.4) |  |

Abbreviations: BAU, Binding Antibody Units
<sup>1</sup>Student t-test or ANOVA test of difference between log transformed geometric mean fold rise; p<0.05 defined as statistically significant difference between groups.
<sup>2</sup>Self reported presence of a serious chronic medical condition such as heart disease, lung disease, diabetes, cancer, liver or kidney disease, immune suppression, or high blood pressure. Missing n=5.
<sup>3</sup>Seropositivity (nucleocapsid BAU/mL $\geq 6.9$ ) based on conversion from mean fluorescence intensity ratio $\geq 1.2$ of sample to calibrator serum in multi-bead assay to BAU/mL using WHO standard serum panel.

Among 7 (7%) patients with ≥1 electronic health record documented prior COVID-19 positive laboratory PCR test, median time since most recent positive COVID-19 test was 351 days (range: 64-758). Among 101 patients who had received at least two doses of COVID-19 mRNA vaccine, median time since receipt of most recent mRNA vaccine dose was 175 days (range: 33-382); 11 had received a booster dose <90 days prior to their enrollment in the study (measured between the date of booster and date of illness onset).

Anti-N and anti-RBD geometric mean fold increases were not associated with days from symptom onset to specimen collection (data not shown). For both antigens, geometric mean fold rise from acute- to convalescent-phase bAb concentrations were similar by patient age, sex, or presence of underlying medical conditions.

In acute-phase specimens, anti-N bAb concentrations were low (GMC: 5.5 BAU/mL, 95% CI: 4.3–7.1; Figure); 72 (69%) participants had anti-N bAb concentrations below the seropositivity threshold of 6.9 BAU/mL. From enrollment to follow-up, anti-N bAb concentration increased 47-fold (CI: 34–66), with convalescent-phase GMC of 259 BAU/mL (CI: 201–335). Among participants with anti-N bAb levels below 6.9 BAU/mL at enrollment, concentrations increased 80-fold (CI: 59–109), versus 15-fold (CI: 7–30) among participants with acute-phase concentrations ≥ 6.9 BAU/mL (*p*-value=0.007). Among four unvaccinated case patients, anti-N bAb concentrations increased 116-fold (95% CI: 7–1998). Patients with prior documented positive COVID-19 tests trended towards having higher convalescent-phase anti-N bAb concentrations than patients without prior positive COVID-19 tests (GMC: 983 [95% CI:546-1771] vs 236 [95% CI: 181-307].

**Figure:**
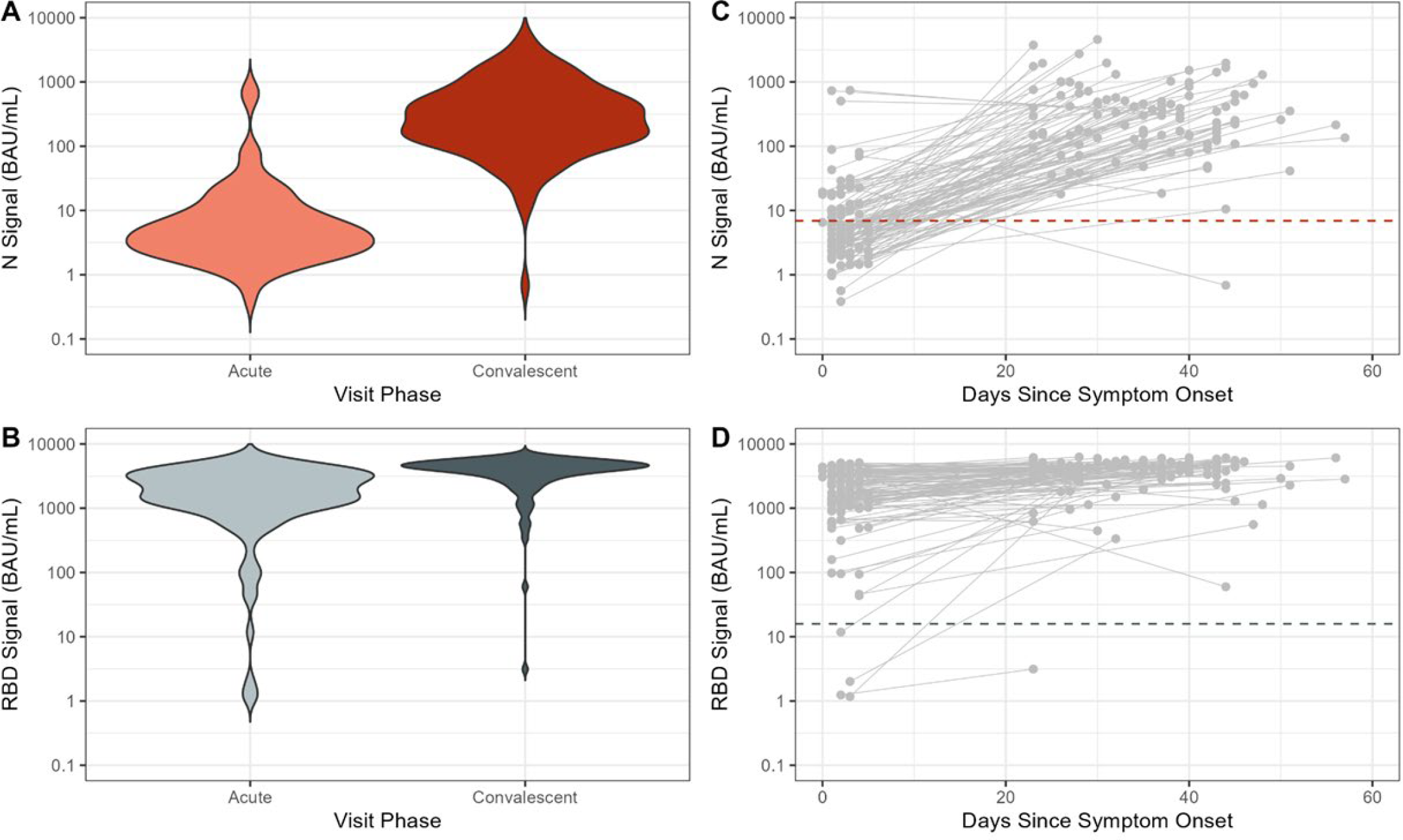
Concentrations of antibodies against SARS-CoV-2 nucleocapsid (N) and ancestral spike protein receptor binding domain (RBD) antigens during acute and convalescent phases of symptomatic COVID-19 associated with Omicron variant virus infection, December 2021—June 2022. (**A**) Antibody concentrations (in binding antibody units [BAU]/mL) against SARS-CoV-2 nucleocapsid protein (N) and (**B)** ancestral spike protein receptor binding domain (RBD) observed in acute and convalescent-phase dried blood spot specimens collected from individuals with symptomatic COVID-19; (**C**) Changes in N concentration and (**D**) RBD concentration in days since symptom onset between acute and convalescent specimens from individuals with symptomatic COVID-19; acute and convalescent-phase specimens from the same individual (N=105) are connected by solid gray line.

Geometric mean anti-spike RBD bAb concentrations were 1258 (CI: 924–1712) and 3189 (CI: 2639–3853) in acute- and convalescent phase specimens, respectively; mean fold RBD bAb rise was 2.5 (CI: 1.9–3.3; Table; Figure). Geometric mean fold rise in RBD bAb was higher among patients who had received 2 mRNA vaccine doses (7.9 [CI: 2.6—24.3]) versus ≥3 doses (2.0 [CI: 1.6—2.5]). Acute- and convalescent-phase anti-RBD bAb levels were low among four unvaccinated case patients (GMC: 16.0 [CI: 0.2–1557] acute and 149.0 [CI: 0.3—273.1] convalescent).

## DISCUSSION

Among symptomatic patients infected with SARS-CoV-2 Omicron variant viruses, most of whom had been previously vaccinated, we observed marked differences in antibody levels against nucleocapsid protein and ancestral spike RBD antigen measured during acute illness and convalescence. The 47-fold increase in IgG antibodies against SARS-CoV-2 nucleocapsid protein indicated a strong humoral response to infection with Omicron variant viruses, mainly BA.1 and BA.2. Baseline seronegative patients had a higher mean fold rise in anti-nucleocapsid antibodies than seropositive patients. Increased convalescent anti-N bAb levels among baseline seropositive patients suggests boosting of immune responses acquired from past infection; this boosting was also observed among patients with ≥1 documented prior COVID-19 positive test. In contrast, we observed modest 2.5-fold increase in antibodies against ancestral spike RBD antigens. Among vaccinated cases, lower baseline anti-RBD antibody concentrations among patients who had received 2 versus ≥3 mRNA vaccine doses were associated with greater antibody response but similar convalescent antibody concentrations. While anti-RBD bAb levels correlate with protection from infection with pre-Omicron variants [1, 3, 4], antibody levels measured against Omicron variants are greatly reduced [8, 14].

Dried blood spots for SARS-CoV-2 serology were collected as part of a COVID-19 vaccine effectiveness study using a test-negative design. Because antibody levels measured during acute illness reflect an estimation of antibody levels close to the time of infection rather than response to current infection, comparison between antibody levels among case patients and SARS-CoV-2 uninfected patients may provide a measure of correlates of risk [15]. However, data, though limited, suggests that vaccination status and pre-existing neutralizing antibodies may affect anti-N antibody response by altering viral load in early illness [8, 16]. Measurement of immune response to infection and antibody levels after convalescence could improve understanding of vaccinated cases and hybrid immunity [17].

The study had several limitations. The serologic assay used in our study contained ancestral SARS-CoV-2 antigens and serum was quantified using the WHO international serum standards from early in the COVID-19 response. Against pre-Omicron variants, virus neutralization titers and IgG antibody concentrations were associated with protection; however, antibody binding and neutralization activity was lower against Omicron variants [2, 8, 18]. In addition, anti-N bAb seropositivity cut-off values were based on mean fluorescence intensity using serum standards rather than blood spots [10, 13]. All case patients included in this study had mild illness; baseline antibody levels and immune responses may differ among patients with severe or prolonged SARS-CoV-2 infection [19]. Among patients with past infection, initial anti-N bAb concentrations may have waned below the seropositivity thresholds [20, 21]. Finally, this analysis included only four unvaccinated case patients, limiting ability to compare unvaccinated infections and reinfections with vaccine breakthrough infections.

Serologic assays that quantify anti-SARS-CoV-2 specific antibody levels during and following acute infection may provide information on incidence and recency of infection. As new SARS-CoV-2 variants emerge, frequent updates to serologic antigens will be needed to quantify binding IgG antibody levels that correlate with immune protection [14]. Observational studies will be critical for evaluating immune responses to COVID-19 vaccination and infection.

## Data Availability

All data produced in the present study are available upon reasonable request to the authors

## Acknowledgments

We acknowledge investigators, study staff and participants from study sites, investigators from the Influenza Division and Malaria Branch, CDC.

## Declarations

Dr. Zimmerman reports grants from CDC, during the conduct of the study, and grants from Sanofi Pasteur, outside the submitted work. Dr. Grijalva reports other from CDC, grants from NIH, other from FDA, grants and other from AHRQ, other from Merck, and other from Syneos Health, outside the submitted work. Dr. Talbot reports grants from CDC, during the conduct of the study. All other authors report not conflicts of interest.

## Disclaimer

The findings and conclusions in this report are those of the authors and do not necessarily represent the official position of the Centers for Disease Control and Prevention. All authors have reviewed and approved of this version of the manuscript.

